# Validation of Machine Learning Models for classification of bladder dysfunction and risk of hydronephrosis in patients with Spina Bifida using videourodynamic data

**DOI:** 10.64898/2026.09.30.26364423

**Authors:** John K. Weaver, Joseph Logan, Julie Klock, Kristina Dortche, Hassaan A. Bukhari, David Buckinsky, Madison Burns

## Abstract

**Purpose:** Videourodynamics (VUDS) are used to assess bladder dysfunction and risk of upper urinary tract deterioration in patients with spina bifida (SB), but interpretation is subject to interrater variability. Machine learning models have previously classified bladder dysfunction severity and predicted incident hydronephrosis using VUDS data. This study evaluated the performance of adapted versions of these models in an independent external cohort.

**Materials and Methods:** VUDS data were collected from SB patients who underwent VUDS at a single institution between 2016 and 2025. Previously developed machine learning frameworks were adapted for implementation in the independent dataset. Three models were utilized: a deep learning convolutional neural network model using pressure-volume data, a deep learning imaging model using fluoroscopic imaging data, and an ensemble model that averaged the risk data from the pressure-volume and fluoroscopic data. The models were evaluated for prediction of incident hydronephrosis and classification of bladder dysfunction severity compared to expert pediatric urologist reviewers.

**Results:** 70 patients were included in the hydronephrosis cohort, of whom 13 (18%) developed incident hydronephrosis. The ensemble model outperformed individual modalities, with a concordance index of 0.77 (pressure-volume only: 0.70, imaging only: 0.72) and an overall AUROC of 0.75 (pressure-volume: 0.67, imaging: 0.74). For bladder dysfunction classification in 95 VUDS studies, the ensemble model achieved 71% accuracy compared to 60% for pressure-volume data alone and 66% for imaging alone, with no substantial disagreements with expert reviewers.

**Conclusions:** The machine learning models that were developed to predict incident hydronephrosis and classify the severity of bladder dysfunction using VUDS data were able to be adapted to an external, independent dataset and perform with similar discrimination and accuracy as previously reported.

## Introduction

Spina bifida (SB) is a congenital neural tube defect impacting 3-4 per 10,000 live births in developed countries^1^. Urologic dysfunction is common in this condition, with 24-98% of patients developing neurogenic bladders^2,3^. Neurologic pathway disruptions can result in detrusor over- or underactivity, incomplete bladder emptying, and detrusor-sphincter dyssynergia (DSD), leading to progressive loss of bladder compliance^3^. Reduced compliance and the resulting elevated bladder pressures increase the risk of hydronephrosis and renal injury, especially in those with neurogenic bladders. Approximately 50% of SB patients will develop kidney disease^2,4–7^.

Videourodyamic studies (VUDS) is the primary tool to evaluate bladder and urethral function and estimate future risk of kidney disease. Approximately 15 – 30% of children with SB have high-risk bladder characteristics seen during VUDS, including presence of DSD, a detrusor leak point pressure (DLPP) > 40 cm H2O, impaired compliance (end filling pressure < 20 mL/cm H2O), a contracted or trabeculated bladder, or vesicoureteral reflux (VUR)^8,9^. Despite the importance and wide use of this study, VUDS interpretation is subject to substantial interobserver variability, which may lead to inconsistent identification of high-risk bladders. In one study, 14 pediatric urologists demonstrated only fair-to-moderate agreement when assessing bladder capacity, end-filling detrusor pressure, detrusor overactivity, and EMG synergy^10^.

Machine learning can offer more objective and reproducible interpretation of VUDS findings. Our team developed machine learning models to classify bladder dysfunction compared to expert reviewers and to predict incident hydronephrosis^11,12^. These models were developed using VUDS data from 306 and 554 patients with SB at a single institution, respectively. The models developed in these studies demonstrated good discrimination when classifying bladder dysfunction and predicting incident hydronephrosis. While internal validation was used to evaluate predictive performance on new data, it may overestimate model performance, necessitating external validation on an independent cohort to assess generalizability.

In this study, we sought to externally validate our previously developed models. We hypothesized that model performance for classifying bladder dysfunction and predicting incident hydronephrosis would be comparable in the external validation cohort to that observed in the original internal cohort.

## Methods

### Study Design and Cohort

Following Institutional Review Board approval, we performed a retrospective cohort study of spinda bifida patients who underwent VUDS at the Cleveland Clinic between 2016 and 2025. Patients were excluded if they underwent surgical intervention that substantially altered bladder storage or emptying physiology (i.e. vesicostomy, bladder augmentation) or if they had a history of imperforate anus.

### Outcomes

This study was designed to evaluate the performance of previously described machine learning models for interpreting VUDS, adapted for implementation in an independent, external institutional cohort. Two outcomes were evaluated: 1) predicting incident hydronephrosis following VUDS and 2) classifying the severity of bladder dysfunction compared to expert review.

#### Incident Hydronephrosis

The primary outcome was incident hydronephrosis on ultrasound performed after the index VUDS. Hydronephrosis was used as a surrogate for the progression of kidney disease, as upper tract dilation is associated with renal tract deterioration in patients with neurogenic bladder^13^.

The presence or absence of hydronephrosis was determined from clinical radiology reports and confirmed by urologist review (JKW) of the ultrasound. Hydronephrosis was considered present if any of the following criteria were identified: 1) ≥5 mm anterior-posterior diameter of renal pelvis on transverse imaging, 2) any major or minor calyceal dilation, or 3) ureteral dilation ≥ 1 mm on supine imaging. For the analysis of incident hydronephrosis, the index VUDS served as time zero. Patients were followed from the index study until the development of hydronephrosis or until their most recent ultrasound.

#### Bladder Dysfunction Severity

The secondary outcome was determining the severity of bladder dysfunction, categorized as a three-level ordinal variable: none/mild, moderate, and severe. The ground truth was defined by three expert reviewers, all of whom were fellowship-trained pediatric urologists who regularly care for patients with SB. Reviewers were provided with the tracings and fluoroscopic images from the VUDS. The reviewers were blinded to all other patient data, such as future renal injury. Classification was based on the overall perceived risk of upper urinary tract dysfunction, with increasing severity corresponding to increasing risk of renal injury.

The final bladder dysfunction severity label for each study was determined by the majority classification of the three expert reviewers.

### Videourodynamic Studies

All VUDS were completed on a Laborie machine (Version 14 Rel 10 Build 5A). Fluoroscopic images were obtained in real time via a C-arm connected to the Laborie software. A rectal catheter and patch electromyography were used for all studies. Studies with technical issues (crying baby through the study, poorly functioning equipment) were excluded from analysis. An attending urologist controlled the C-arm during all studies and fluoroscopic images were obtained at 25%, 50%, and 75% of EBC and at times deemed important by the performing urologist.

## I. Urodynamic Tracing Preprocessing

Urodynamic tracing data were obtained from fluoroscopic studies and stored as tab-delimited text files, each corresponding to a single study. Each file contained time-series measurements including intravesical water volume (VH_2_O) and detrusor pressure (Pdet). To normalize across patients with varying bladder capacities, VH_2_O was divided by the patient’s expected bladder capacity (EBC), yielding a percent-full metric ranging from 0 to 1. The percent-full values were rounded to the nearest hundredth, and Pdet measurements were averaged within each percent-full bin using a groupby aggregation, producing a fixed-resolution pressure-over-filling curve per study. The resulting per-study curve was transposed into a single row vector, with each column representing the mean detrusor pressure at a given percent of bladder capacity. In cases where studies did not record measurements across the full filling range, missing values were forward-filled to ensure uniform feature vector length across all subjects. Study identifiers were retained as the first column of the final feature matrix. Processed tracings were saved as a single CSV file for downstream modeling.

## II. 1D CNN Classification of Urodynamic Tracings

Following preprocessing, forward filling was applied along the feature axis to propagate the last observed detrusor pressure value forward into any missing percent-full bins, ensuring each study’s feature vector spanned the complete filling range from 1% to 100% of EBC.The resulting feature matrix was used to train a one-dimensional convolutional neural network (1D-CNN) for three-class severity classification. The target label was the average urodynamic rating assigned by clinician review. Labels were ordinally encoded and class weights of 1.2, 0.7, and 1.4 were applied to classes 1, 2, and 3 respectively to address class imbalance. The 1D-CNN architecture consisted of three successive convolutional blocks, each comprising a Conv1D layer with 64 filters and kernel size 3, followed by batch normalization and ReLU activation. A global average pooling layer aggregated the temporal features into a fixed-length representation, which was passed to a fully connected output layer. The model was trained using cross-entropy loss with Adam optimization at a learning rate of 1×10^−3^ and a batch size of 32 for 20 epochs.

Model performance was estimated using 5-fold stratified cross-validation with a fixed random seed. At each fold a fresh model was instantiated and trained on the training partition, and out-of-fold (OOF) predictions were collected on the held-out validation partition. Predicted class probabilities were written back to their original row positions in the full dataset, such that after all folds every sample had exactly one set of OOF probabilities derived from a model that had not seen that sample during training. Overall performance was reported as mean accuracy across folds alongside a classification report and confusion matrix computed on the aggregated OOF predictions.

## III. Fluoroscopic Image Preprocessing

Fluoroscopic DICOM images were retrieved and matched to study metadata using subject identifiers and filenames. DICOM files were read using pydicom, and pixel arrays were extracted and cast to 32-bit float. Multi-frame DICOM files, where an entire image series was stored as a single file, were handled by selecting the middle frame to obtain a representative single image. Each image was min-max normalized to the range [0, 1] using per-image minimum and maximum values with a small epsilon for numerical stability. Images were resized to 224×224 pixels using OpenCV. To characterize image quality, adaptive thresholding was applied using a mean-based method with a block size of 7 and constant of 13 to produce a binary segmentation mask. External contours were extracted and the largest contour area was recorded as a quality metric alongside per-image mean and median pixel intensity. Images that did not conform to the target resolution after resizing were excluded. Grayscale images were replicated across three channels to produce RGB-compatible tensors. The final image array was saved as a NumPy array and subject-level metadata including source directory, contour area, and pixel statistics were saved to a CSV for downstream quality control and merging.

## IV. ResNet-50 Classification of Fluoroscopic Images

The preprocessed image array and corresponding metadata were loaded for model training. Labels were ordinally encoded from the three-class average urodynamic rating (1, 2, 3) to zero-indexed integers (0, 1, 2). A ResNet-50 architecture pretrained on ImageNet was used as the image encoder, with the final fully connected layer replaced by a linear layer with three output units. Training augmentations included random horizontal flip, random vertical flip, and random rotation up to 15 degrees, followed by ImageNet mean and standard deviation normalization. Validation images received normalization only.

Model performance was evaluated using 5-fold subject-level cross-validation implemented with GroupKFold, ensuring that all images from a given subject appeared exclusively in either the training or validation partition within each fold, preventing data leakage across the patient boundary. Class weights of 1.2, 0.7, and 1.4 were applied to classes 0, 1, and 2 respectively via weighted cross-entropy loss to address class imbalance. Models were optimized using Adam at a learning rate of 1×10^−4^ with cosine annealing over 20 epochs and a batch size of 8. The best-performing checkpoint per fold was selected by validation accuracy, and out-of-fold softmax probabilities were written back to the original frame-level dataframe at their original row positions for downstream ensemble modeling.

### Model Evaluation

Adapted machine-learning models performance was evaluated separately for the two prespecified outcomes. To classify bladder dysfunction severity, overall accuracy and weighted Cohen’s kappa were calculated. Confusion matrices were constructed to characterize patterns of disagreement. Receiver operating characteristic analyses and corresponding areas under the curve with 95% confidence intervals were calculated as appropriate. For prediction of hydronephrosis, model discrimination was assessed within a time-to-event framework. Model discrimination was quantified using the concordance index, with time-dependent receiver operating characteristic curves and area under the curve estimates at prespecified follow-up intervals. Calibration was assessed by comparing predicted and observed probabilities of hydronephrosis. For studies that did not reach 75% of the patient’s estimated bladder capacity (EBC), imputation was used as a proxy. Imputation assignment differentiates this model to prior models that categorized all patients based on EBC.

### Statistical Analysis

Continuous variables were summarized as medians with interquartile ranges. Summary statistics were based on the data distribution and were expressed as median (interquartile range [IQR]) or frequency (percentage).

Hydronephrosis-free survival was estimated using Kaplan-Meier methods where appropriate.

Differences between severity categories (none/mild, moderate, severe) were calculated using the Kruskal-Wallis rank sum test for continuous variables and chi-square for categorical variables.

Statistical tests were two-sided, and a p-value <0.05 was considered statistically significant. Receiver Operating Characteristic Curve (ROC) and Area Under the Curve (AUC) were generated for each subgroup analysis. Using the 75% EBC ensemble model, patients were stratified into high and average risk groups. The results of this stratification using various risk score cutpoints are shown in Supplemental Table 3.

## Results

### Machine Learning Model assessing risk for developing hydronephrosis

70 patients met inclusion criteria. Patients ranged in age from 2 months to 63 years at the time of VUDS. Median age was 175.03 months (IQR 30-404). 54% of patients were under 18 at the time of the study. Demographic characteristics are shown in Table 1. 13 of the 70 patients developed hydronephrosis (18%). The ensemble model performed best overall and by year, with a C-index of 0.765 and overall AUC of 0.750 (Table 2). Risk scores from the ensemble model were used to classify patients as high- or average-risk. Results of stratification by risk score are shown in Table 3. Figure 1 shows the Kaplan-Meier curve for the development of incident hydronephrosis among the top 20% of patients by risk, compared with the rest of the cohort.

**Table 1.**
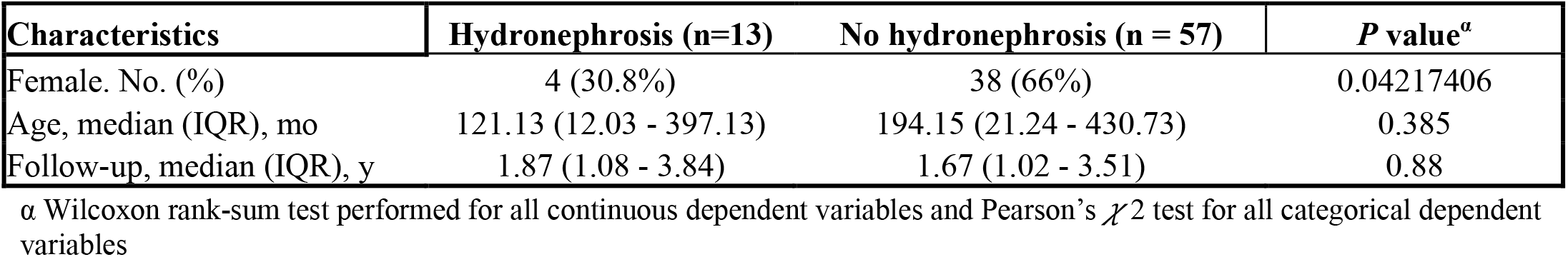
Characteristics of patients.

**Table 2.**
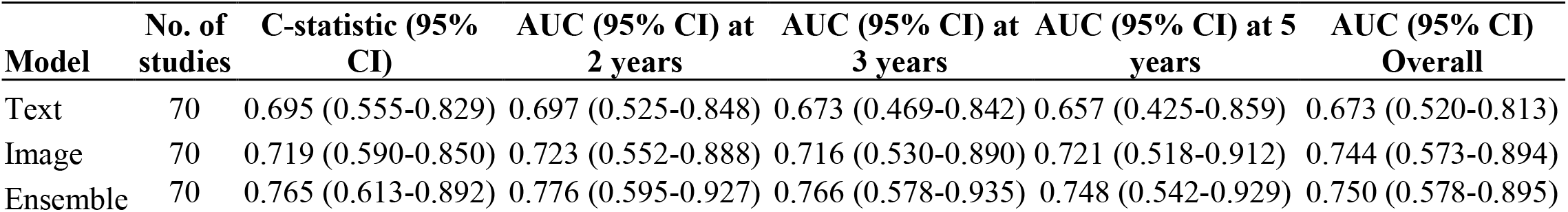
Discrimination of models by year and overall.

**Table 3.**
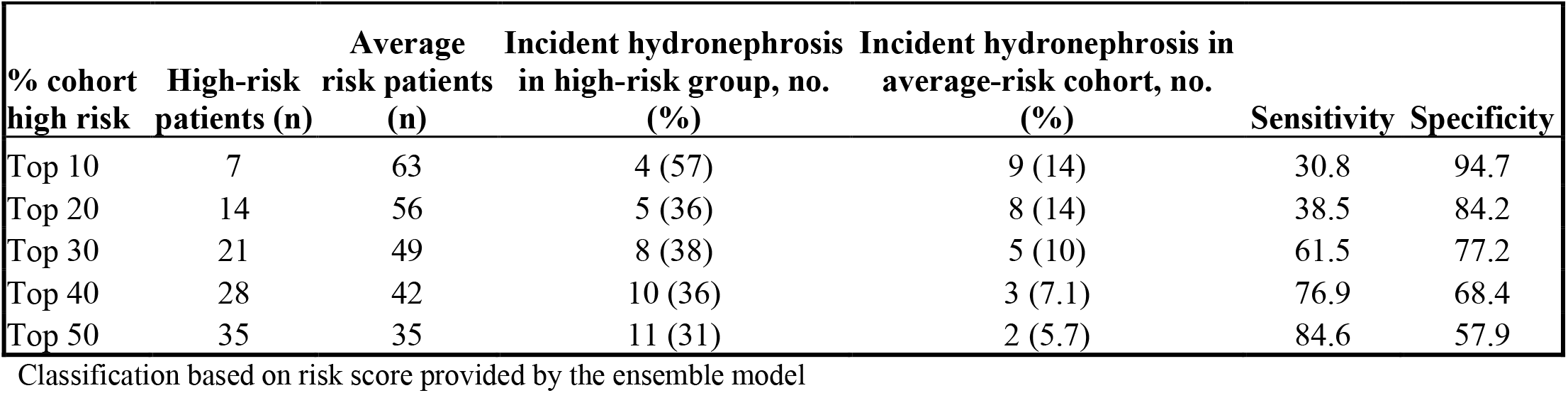
Risk stratification of the cohort.

| % cohort high risk | High-risk patients (n) | Average risk patients (n) | Incident hydronephrosis in high-risk group, no. (%) | Incident hydronephrosis in average-risk cohort, no. (%) | Sensitivity | Specificity |
| --- | --- | --- | --- | --- | --- | --- |
| Top 10 | 7 | 63 | 4 (57) | 9 (14) | 30.8 | 94.7 |
| Top 20 | 14 | 56 | 5 (36) | 8 (14) | 38.5 | 84.2 |
| Top 30 | 21 | 49 | 8 (38) | 5 (10) | 61.5 | 77.2 |
| Top 40 | 28 | 42 | 10 (36) | 3 (7.1) | 76.9 | 68.4 |
| Top 50 | 35 | 35 | 11 (31) | 2 (5.7) | 84.6 | 57.9 |

Classification based on risk score provided by the ensemble model

**Figure 1.**
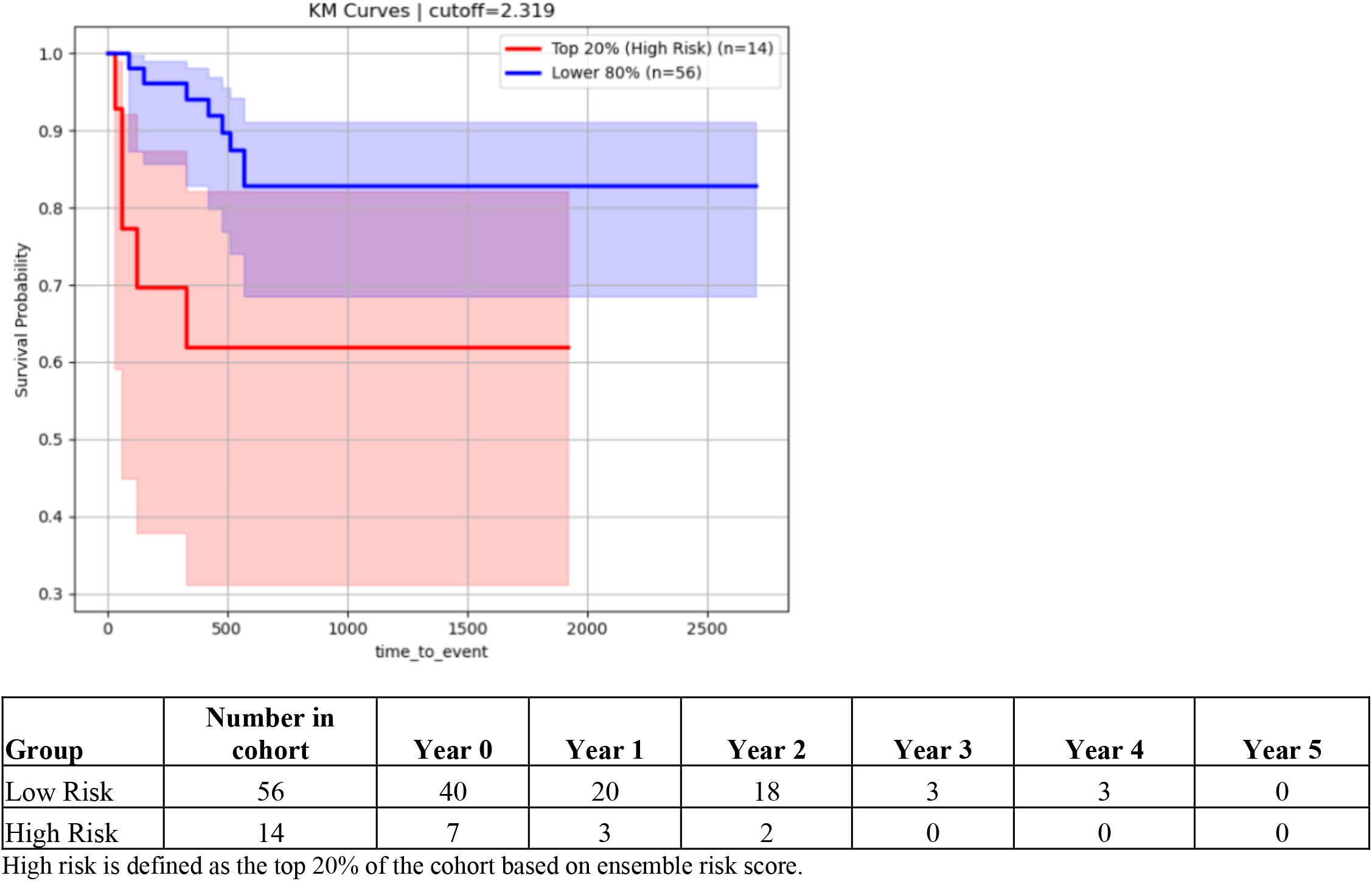
Survival Analysis of incident hydronephrosis between high- and low-risk patients.

### Machine Learning Model classification of bladder dysfunction compared to expert reviewers

70 studies were included for hydronephrosis (Table 2); however, 95 VUDS were included in the analysis due to repeated patients (Table 4 and 5). The ensemble model was the most accurate overall, achieving 71% accuracy and showing no substantial disagreements relative to expert review (Table 4). The image model was the only model to display substantial disagreement. See Table 5 for the model’s performance for each severity rating, compared with expert review, as measured by area under the curve. Table 6 shows the model’s classification compared with that of expert reviewers.

**Table 4.** Performance results of models compared to expert reviewer’s majority ratings.

| Model | Number of studies | Accuracy (95% CI) | Weighted Kappa score | Substantial Disagreement* |
| --- | --- | --- | --- | --- |
| Text | 95 | 59.9% (49.5%, 69.5%) | 0.374 | 0 |
| Image | 95 | 65.6% (55.8%, 74.7%) | 0.394 | 4 |
| Ensemble | 95 | 70.8% (61%, 80%) | 0.492 | 0 |

\*study rated no/mild by experts but severe by model or vice versa

**Table 5.** Performance results of models compared to expert reviewer’s majority ratings using AUC measurements.

| Model | Number of studies | AUC (95% CI) none/mild vs remaining cohort | AUC (95% CI) moderate vs remaining cohort | AUC (95% CI) severe vs remaining cohort |
| --- | --- | --- | --- | --- |
| Text | 95 | 0.78 | 0.64 | 0.88 |
| Image | 95 | 0.73 | 0.68 | 0.7 |
| Ensemble | 95 | 0.81 | 0.72 | 0.86 |

**Table 6.**
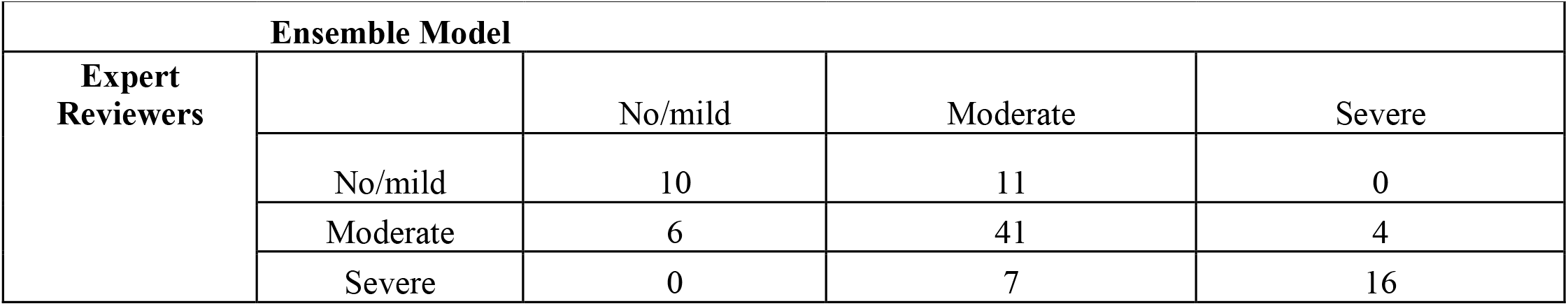
Ensemble model predictions compared to expert reviewer’s majority ratings.

| Ensemble Model |  |  |  |  |
| --- | --- | --- | --- | --- |
| Expert Reviewers |  | No/mild | Moderate | Severe |
|  | No/mild | 10 | 11 | 0 |
|  | Moderate | 6 | 41 | 4 |
|  | Severe | 0 | 7 | 16 |

## Discussion

The previously developed machine learning frameworks were successfully adapted to an independent external cohort while maintaining performance in predicting incident hydronephrosis and classifying bladder dysfunction. Forward-fill imputation was implemented for studies that did not reach 75% of the estimated bladder capacity, broadening the applicability of the models to incomplete VUDS. The ensemble model performed best at predicting incident hydronephrosis and classifying the severity of bladder dysfunction. Utilizing pressure-volume and fluoroscopic data appears to retain predictive value across institutions, despite differences in data acquisition and implementation.

The model’s ability to predict incident hydronephrosis appears to be preserved across institutions. This study’s concordance index of 0.765 is comparable to the best-performing ensemble model in the prior study, which has a concordance index of 0.71^12^. Consistent with the prior study, the ensemble model successfully stratified patients according to risk of incident hydronephrosis in our independent cohort. These findings suggest a potential role for machine learning-based risk stratification in identifying patients at greatest risk for upper tract deterioration who may benefit from closer surveillance and/or earlier intervention.

Classification of bladder dysfunction severity was similar in our cohort compared to the prior study, with previously reported 70% accuracy and weighted κ 0.54 with the 75% EBC ensemble model, compared to the 70.8% accuracy and *k* 0.492 in the ensemble model for our cohort^11^. Notably, there were no substantial disagreements in our cohort. This suggests that machine learning models may be a useful adjunct to VUDS interpretation, especially given the known interrater variability in this domain^10^.

Our study has several limitations. Our cohort was relatively small, with only 13 hydronephrosis events. Future studies would benefit from implementing these models with larger datasets. Incident hydronephrosis was also used as a proxy for upper-tract deterioration rather than for the actual progression of kidney disease. Additionally, the rating of bladder dysfunction severity was based on expert reviewers rather than objective biological reference standards, despite interrater variability. Our study is also uniquely limited by the differences in VUDS data acquisition between the original institution and ours. For this reason, the machine learning framework had to be adapted. One such adaptation was the forward-fill imputation for the studies that did not reach 75% of the EBC. While this broadens applicability, it also introduces assumptions about the pressure-volume trajectory after study termination. Prospective and retrospective multicenter studies are needed for validation, including those evaluating the model’s performance in patients with non-SB neurogenic bladder.

## Conclusion

Machine learning models for VUDS interpretation maintained predictive and classification performance when adapted to an independent external cohort. These findings support further multicenter evaluation of these models as tools for risk stratification in patients with SB.

## Data Availability

All data produced in the present study are available upon reasonable request to the authors

